# HGACL-DRP: Heterogeneous Graph Attention Dual-Perturbation Contrastive Learning Network for Drug Response Prediction

**DOI:** 10.1101/2025.09.22.25336318

**Authors:** Min Li, Ming Jin, Shaobo Deng

## Abstract

The marked heterogeneity of cancer poses a substantial challenge to precision drug therapy, resulting in considerable variability in patient responses to identical treatments. Accurately predicting drug sensitivity thus represents an urgent and critical challenge in the field of pharmacology and personalized medicine. Existing methods have limitations in addressing the complexity of multi-modal biological data and heterogeneous graphs. Additionally, their heavy reliance on labeled data hinders model generalization. To address these challenges, we propose the Heterogeneous Graph Attention Dual-Perturbation Contrastive Learning Network for Drug Response Prediction (HGACL-DRP). This framework constructs an optimized heterogeneous graph structure by integrating multi-omics features and implementing a robust neighbor filtering mechanism. HGACL-DRP innovatively introduces a Dual-Perturbation contrastive learning paradigm that incorporates both feature and structural perturbations specifically designed for drug response prediction, thereby enabling the self-supervised learning of more robust heterogeneous graph node embeddings. On two major public datasets, the Genomics of Drug Sensitivity in Cancer (GDSC) and the Cancer Cell Line Encyclopedia (CCLE), HGACL-DRP exhibits exceptional performance across multiple classification metrics, achieving a mean AUC of 95.48% on CCLE and 98.99% on GDSC. Compared to existing state-of-the-art drug response prediction algorithms, HGACL-DRP demonstrates statistically significant improvements in predictive accuracy. The innovative multi-view graph augmentation strategy and attention mechanism are designed to enhance personalized drug selection and facilitate novel drug discovery. HGACL-DRP is anticipated to address the challenges posed by drug response heterogeneity, thereby providing reliable support for clinical decision-making.

## 1. Introduction

Cancer, a global health challenge, presents a significant threat to human well-being due to its profound heterogeneity. This inherent variability means that patients diagnosed with the same type of cancer may exhibit vastly different clinical responses to identical drug therapies [1]. Consequently, this inherent heterogeneity renders the development of personalized and effective drug treatment regimens for each patient a central challenge and an urgent priority in the field of precision medicine [2].

In recent years, the advent of high-throughput sequencing technologies and large-scale drug screenings has led to the rapid accumulation of comprehensive public databases, such as the Cancer Cell Line Encyclopedia (CCLE) and the Genomics of Drug Sensitivity in Cancer (GDSC). These databases serve as critical resources, offering a solid foundation for computational methods focused on predicting anti-cancer drug response. By leveraging this kind of data, researchers have developed diverse computational models designed to extract features from cell lines and drugs, predict their interactions, and ultimately guide drug discovery and personalized medicine [3].

Existing Drug Response Prediction (DRP) methods differ in their strategies for feature extraction and model construction. Early studies predominantly concentrated on extracting features from cell line omics data and drug chemical structures, which were subsequently utilized as inputs for traditional machine learning models to perform predictions. For example, Su et al. transformed features such as gene expression and copy number variation into high-dimensional vectors through multi-grained scanning and employed a cascade forest model for classification [4]. Sharifi-Noghabi et al. leveraged deep neural networks to learn representations from various omics data types, including somatic mutations, copy number variations, and gene expression, before concatenating these learned features for drug response prediction [5]. Shin et al. embedded knowledge-guided GNNs within a Transformer framework to predict drug responses from gene pathways and chemical structures, yielding more interpretable predictions [6]. Liu et al. achieved interpretable predictions by decomposing drugs and cell lines into sub-components to pinpoint the most influential drivers of response outcomes [7]. Despite advancing the field of DRP to some extent, these methods are often limited by challenges in effective feature extraction and a relatively restricted ability to model non-linear relationships.

With the emergence of Graph Neural Networks (GNNs) for processing complex relational data, they have demonstrated substantial potential in uncovering relationships between drugs and cell lines. A growing body of research has focused on representing drugs and cell lines as graph structures to harness the robust feature extraction capabilities of GNNs[8]. These approaches typically involve constructing a heterogeneous network that integrates cell lines, drugs, and their known interactions. MOFGCN [9] stands out as a representative study that first integrates multi-omics data (gene expression, copy number variation, somatic mutation) to compute cell line similarity, subsequently builds a heterogeneous network by incorporating drug fingerprint similarity and known cell line-drug associations, and finally extracts latent features through graph convolution operations. Building on this, NIHGCN [10] introduced a parallel heterogeneous graph convolutional network based on Neighborhood Interaction (NI). This approach aims to simultaneously aggregate node-level and element-level neighbor features, thereby enabling more effective capture of the intrinsic differences and interactions between cell line and drug nodes. TSGCNN [11] innovatively constructed separate feature spaces for cell lines and drugs, performing graph convolution within these homogeneous spaces to propagate similarity information. Subsequently, it aggregated features from different node types in the heterogeneous network to generate final representations. Wang et al. introduced a hierarchical attention network that integrates a graph attention mechanism (to capture external relationships between entities) with a multi-head self-attention mechanism (to reinforce internal correlations among features), achieving a fusion of multi-scale relationships to further enhance DRP performance [12].

To address the challenges of data sparsity and limited labels, Contrastive Learning has been adopted in the DRP field as a powerful self-supervised learning paradigm [13]. Liu et al. integrated GNNs with contrastive learning: by contrasting the embeddings of a cancer drug-response graph with those of its corresponding resistance graph, they boosted the model’s discriminative power and generalization capability [14]. Dong et al. implemented an adaptive sparse mapping-based graph contrastive learning network that performs graph augmentation using a GraphMorpher module (featuring node attribute masking and topological pruning) and designed multi-level contrastive learning tasks to enhance feature discrimination [15]. Lee et al. introduced a response-aware multi-task learning framework that combines a Bayesian neural network with soft-supervised contrastive regularization, effectively addressing the data imbalance commonly observed in drug response datasets [16].

Recently, there has been a growing focus in related studies on the application of drug response prediction. For example, DeepCDR [17] is a hybrid graph convolutional network that integrates multi-omics data with drug chemical structures for predictive modeling. Wang et al. proposed a kernel-based deep learning framework that integrates heterogeneous data by constructing kernel similarity matrices derived from multi-source omics data for drug repurposing tasks [18]. Lao et al. utilized sequence recombination-based data augmentation and edge collaborative update strategies to enhance the efficiency of drug molecular feature extraction [19]. Kim et al. innovatively leveraged an adversarial network to minimize the distributional discrepancy between preclinical and clinical datasets, thereby improving the model’s generalization to external clinical data [20].

Despite these advancements, existing DRP methods continue to encounter numerous challenges in drug response prediction. First, while multi-omics datasets offer a more comprehensive understanding of biological processes, effectively integrating this heterogeneous information and modeling the complex, non-linear interactions among different omics types remain significant technical obstacles. Second, traditional graph neural network (GNN) approaches, when constructing heterogeneous graphs, frequently emphasize the associations between heterogeneous entities such as cells and drugs but overlook potential deeper similarities and interactions among homogeneous nodes (e.g., cell-cell or drug-drug), leading to suboptimal information utilization. Moreover, most models are heavily reliant on label-supervised learning; however, high-quality annotated data, such as known drug responses, are often scarce and limited in the biomedical domain, thereby constraining model generalization, particularly for predicting responses in new cell lines or drugs. Lastly, pure prediction outcomes typically lack interpretability, hindering the discovery of underlying drug action mechanisms, which is essential for guiding clinical applications and facilitating novel drug development.

To address these critical challenges, this paper presents a novel Heterogeneous Graph Attention Dual-Perturbation Contrastive Learning Network for Drug Response Prediction (HGACL-DRP), which is specifically designed to achieve precise and robust drug response prediction. This is accomplished by integrating multi-modal omics data, optimizing the graph structure, and leveraging a self-supervised contrastive learning framework tailored for DRP. The primary contributions of this work are as follows:

1. HGACL-DRP introduces an attention-driven multi-modal feature fusion mechanism that leverages distinct kernel methods tailored for each type of omic data and incorporates an attention-based dynamic weighting scheme to enable more flexible and adaptive fusion.
2. HGACL-DRP employs a robust neighbor filtering mechanism guided by node attention scores, which effectively prunes the similarity networks to reduce noise and construct a more streamlined graph structure.
3. HGACL-DRP introduces a novel Dual-Perturbation contrastive self-supervised learning framework to learn more robust node embeddings from sparsely labeled data. This framework generates multiple graph views through structural and feature perturbations and employs a tailored InfoNCE loss for self-supervised training, significantly enhancing the model’s generalization capability and discriminative power.
4. HGACL-DRP is an end-to-end framework that transforms learned heterogeneous graph node embeddings into drug sensitivity predictions through a graph decoder. This approach enables seamless and computationally efficient processing from raw data to final predictions. In comparison with existing state-of-the-art DRP algorithms, HGACL-DRP exhibits substantial performance improvements.

The remainder of this paper is organized as follows: Section 2 elaborates on the experimental data and methodology employed in HGACL-DRP. Section 3 presents the experimental results along with an in-depth analysis. Finally, Section 4 summarizes the key findings and concludes this work.

## 2. Experimental Data and Methodology

This section delineates the empirical foundation of our study, beginning with a thorough overview of the data sources and their rigorous preprocessing. A explanation detailed of the HGACL-DRP architecture follows, highlighting its core components and their collaborative roles in enabling precise drug response prediction. This systematic approach enhances reproducibility and provides clarity in comprehending the methodological framework underpinning our proposed solution.

### 2.1. Experimental Data

This study focuses on model construction and evaluation by leveraging two widely-used public drug sensitivity databases: the Genomics of Drug Sensitivity in Cancer (GDSC) (https://www.cancerrxgene.org/) and the Cancer Cell Line Encyclopedia (CCLE) (https://sites.broadinstitute.org/ccle). These databases provide comprehensive experimental data regarding the responses of multiple cancer cell lines to various anti-cancer drugs, along with associated multi-omics features, thereby establishing a robust foundation for computational research in drug response prediction.

In the GDSC database, we primarily utilized the provided half-maximal inhibitory concentration (IC50) values and drug sensitivity thresholds. Based on these thresholds, we categorized the responses of cell line-drug combinations into binary categories: a cell line was deemed sensitive (labeled as 1) if its IC50 value was less than or equal to the corresponding drug’s sensitivity threshold; otherwise, it was classified as resistant (labeled as 0). After preprocessing, we obtained response data for 962 cell lines across 228 drugs from the GDSC database, comprising 20,851 sensitive samples and 156,512 resistant samples. This binarization is a common practice in computational drug response prediction, which effectively transforms the task from a continuous regression problem into a more manageable binary classification problem. This strategy—declaring a cell line sensitive or resistant—has been widely adopted and empirically validated as an effective way to capture the primary therapeutic outcome of interest [9–11,14,15].

The CCLE database offers an extensive collection of cell line drug trial records, encompassing detailed information such as drug targets, dosages, and logarithmic IC50 values. Similar to GDSC, we evaluated drug sensitivity using the logarithmic IC50 values. Specifically, a cell line was classified as sensitive if its Z-score normalized logarithmic IC50 value was below -0.8; otherwise, it was categorized as resistant. After preprocessing, the CCLE dataset encompassed response data for 436 cell lines across 24 drugs, including 1696 sensitive and 8768 resistant samples.

In addition to drug response data, this study integrates multiple key omics datasets to comprehensively characterize cell lines and drugs. Specifically, we leverage three distinct types of omics data for cell lines: Gene Expression, Copy Number Variation (CNV), and Somatic Mutation, obtained from the GDSC and CCLE databases.

Gene Expression: These data form a matrix in which each entry gives the normalized expression level of a specific gene in a given cell line. Raw RNA-seq counts are log2-transformed and otherwise processed to correct for batch effects.

Copy Number Variation (CNV): CNV is represented as a matrix indicating the number of copies of specific genomic regions. The original data undergo processing to segment the genome and call CNVs for each cell line.

Somatic Mutation: A binary matrix encoding the presence (1) or absence (0) of each somatic mutation in every cell line.

Drug molecular features are encoded as substructure fingerprints retrieved from PubChem. Each fingerprint is a binary vector—essentially a one-hot encoding—that records the presence (1) or absence (0) of predefined chemical fragments, thereby capturing the structural profile of the corresponding drug. All multi-modal features, including both omics and chemical descriptors, are pre-processed and standardized to ensure consistent, high-quality inputs for the model.

During the model training process, missing values in the drug response matrix (i.e., unknown associations) are masked and excluded from the loss calculation to ensure that the model is optimized solely based on known data. Subsequently, the preprocessed cell line omics data, drug fingerprint data, and cell line-drug association data are integrated and utilized as the input for the model.

### 2.2. Methodology

This section provides a detailed description the architecture and key components of our proposed Heterogeneous Graph Attention Dual-Perturbation Contrastive Learning Network (HGACL-DRP). HGACL-DRP is an end-to-end deep learning framework designed to learn robust representations of cell lines and drugs by integrating multi-omics features and drug molecular structures, thereby enabling accurate prediction of drug response.

#### 2.2.1. Overview

As illustrated in **Fig. 1**, the overall framework of HGACL-DRP can be broken down into several key stages: First, the multi-modal feature fusion module conducts kernel extraction and attention-driven fusion on the raw omics data of cell lines and drugs, thereby generating high-quality node feature representations. Second, the heterogeneous graph construction and augmentation module employs a robust neighbor filtering mechanism to refine the graph structure, constructing a heterogeneous graph that incorporates cell-cell similarity, drug-drug similarity, and cell-drug associations. Next, the Dual-Perturbation contrastive self-supervised learning module creates multiple graph views by introducing both feature and structural perturbations, enabling the learning of more discriminative node embeddings through a contrastive learning paradigm. Finally, the drug response prediction module leverages the learned node embeddings to reconstruct the drug response association matrix via a graph decoder, thereby completing the final drug sensitivity prediction task.

**Fig. 1.**
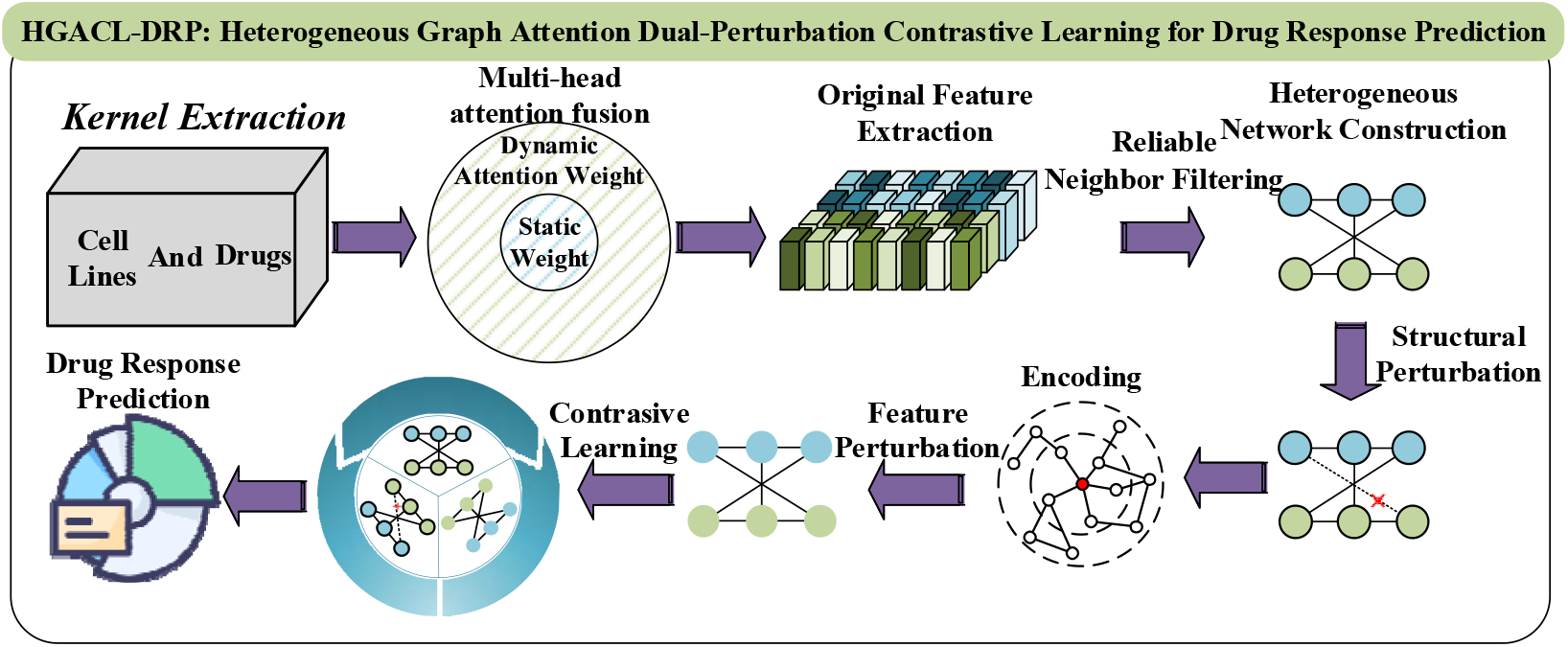
Overall Framework of HGACL-DRP.

#### 2.2.2. Multi-Modal Feature Fusion

This module constitutes the initial stage of HGACL-DRP, with the primary goal of transforming heterogeneous raw data from cell lines and drugs into unified, high-information-content node feature representations capable of capturing complex biological associations. Given the complexity of biological processes, single-omics data often fall short in providing a comprehensive understanding of a cell’s state or a drug’s properties. In contrast, multi-omics data fusion offers a more holistic perspective. Moreover, different omics data inherently exhibit distinct distributions and characteristics (e.g., gene expression is continuous, whereas mutations are discrete). Conventional methods such as simple concatenation or averaging may fail to effectively capture the deep non-linear relationships between these data types and could even introduce noise. To address these challenges, as illustrated in Fig. 2, this module introduces innovations like kernel-based similarity transformations for heterogeneous data alignment and an attention mechanism for adaptive weight fusion, thereby generating more robust and expressive feature representations.

**Fig. 2.**
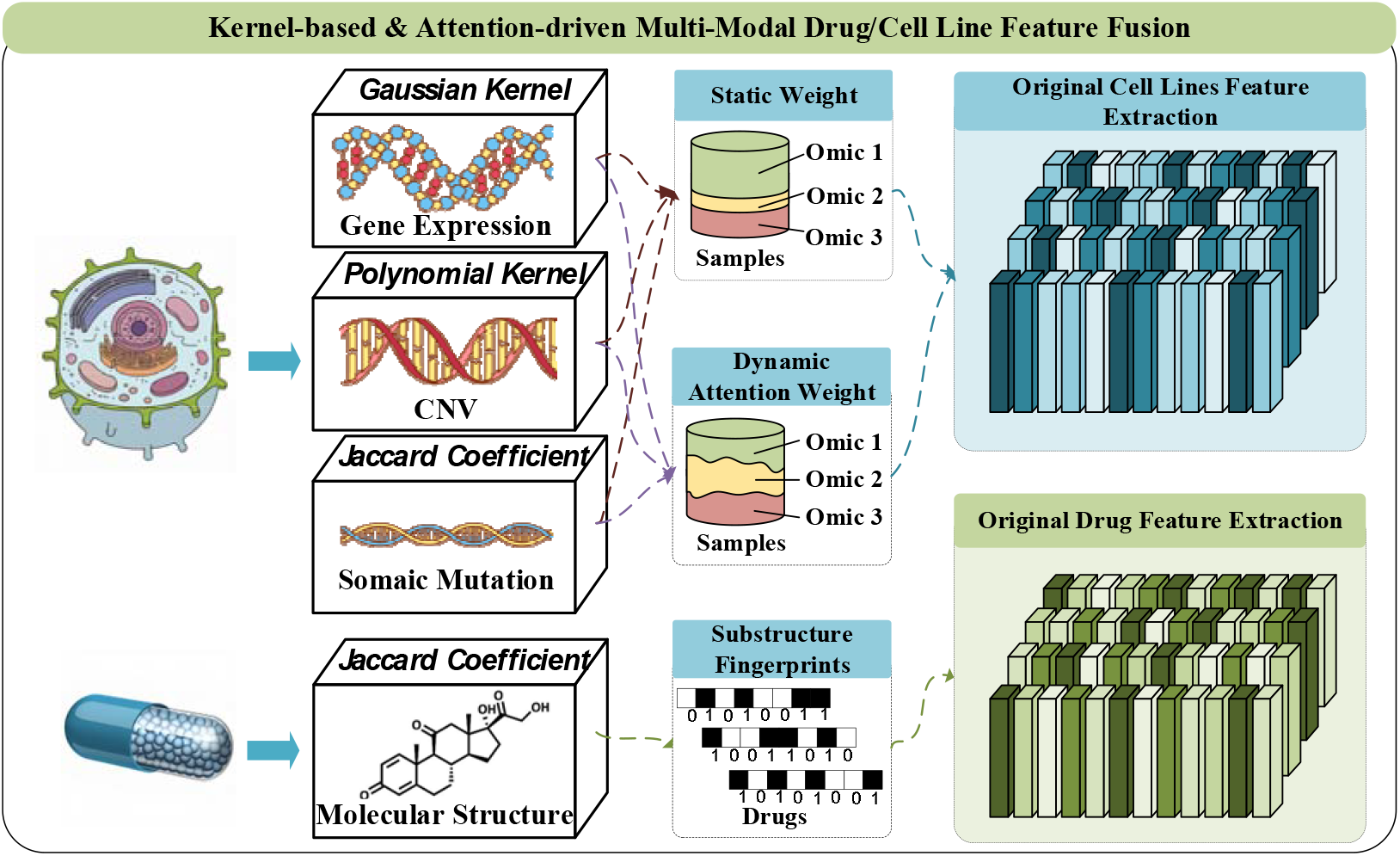
Multi-Modal Feature Fusion.

This module systematically integrates diverse omics and drug features through a two-step process: kernel-based similarity mapping and an attention-driven fusion mechanism. We first generate similarity matrices for each omics type and for drugs, selecting kernels tailored to data characteristics—a Gaussian kernel for continuous data such as gene expression, a Polynomial kernel for non-linear patterns in CNA data, and the Jaccard coefficient for sparse binary data such as mutations and drug fingerprints. Next, a hybrid attention mechanism that combines fixed static weights with learned dynamic weights is applied to these similarity matrices to produce a robust fused representation. The final representation is a weighted sum of the individual kernels, with weights obtained by equally combining the static and dynamic components, ensuring balanced and comprehensive feature integration.

##### Original Feature Embedding

The three omics data types for cell lines—Gene Expression ***F***_***gene*,**_ Copy Number Variation ***F***_***cna***_, and Somatic Mutation ***F***_***mutation***_—along with the drug substructure fingerprints ***F***_***drug***_, are initially mapped to a unified, shared low-dimensional space ***E***_***gene*,**_ ***E***_***cna***_,,***E***_***mutation***_, ***E***_***drug***_ through their respective independent linear transformation layers.

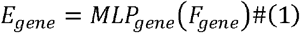

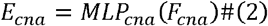

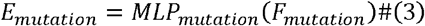

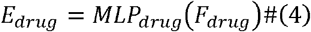

where ***MLP*** represents the linear transformation layers to map these raw features into a unified, shared low-dimensional embedding space of dimension ***d***_***shared***_ (128 in our implementation). This step lays the foundation for subsequent similarity calculations and fusion operations by resolving dimensional mismatches.

To respect the intrinsic characteristics of different omics data, we innovatively incorporate kernel methods to compute their similarities instead of directly fusing raw features. This approach is advantageous as it allows kernel methods to map non-linear relationships in the original feature space into linear relationships in a high-dimensional Hilbert space. Consequently, this enables the capture of more complex non-linear similarities and provides a biologically more meaningful similarity metric for subsequent fusion.

##### Gaussian Kernel for Gene Expression

Gene expression data typically exhibits a continuous distribution, and the similarity between samples can be quantified using Euclidean distance. The Gaussian kernel transforms this distance into a similarity score within the range [0, 1], where smaller distances correspond to higher similarity.

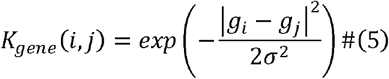

where ***g***_***i***_ and ***g***_***j***_ are the gene expression vectors for cell lines ***i*** and ***j***, respectively, and ***σ*** is the bandwidth parameter of the Gaussian kernel, which controls the rate of similarity decay.

##### Polynomial Kernel for Copy Number Variation

CNA data reflects copy number alterations in genomic segments and may demonstrate strong non-linear associations. The polynomial kernel can effectively capture such non-linear similarities by leveraging a polynomial form.

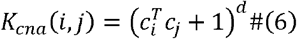

where ***c***_***i***_ and ***c***_***j***_ are the CNA vectors for cell lines ***i*** and ***j***, respectively, and ***d*** is the degree of the polynomial kernel, which controls the degree of non-linearity.

##### Jaccard Coefficient for Somatic Mutation and Drug Fingerprints

Mutation data and drug fingerprints are generally characterized as binary, sparse, and discrete features. The Jaccard coefficient is especially appropriate for measuring the similarity of such binary or set-based data, as it calculates the ratio of the size of the intersection to the size of the union of two sets, thereby effectively capturing the shared characteristics between them.

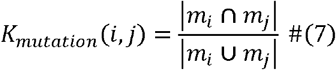

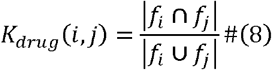

where ***m***_***i***_ and ***m***_***j***_ are the mutation vectors for cell lines ***I*** and ***j***, and ***f***_***i***_ and ***f***_***j***_ are the fingerprint vectors for drugs ***i*** and ***j***. All generated kernel matrices are normalized using the Frobenius norm before subsequent fusion to ensure consistent scaling:

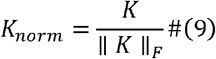

where ∥ · ∥ _***F***_ denotes the Frobenius norm.

##### Attention-Driven Multi-Modal Kernel Fusion

After computing the kernel matrix for each modality, an attention-driven fusion mechanism is introduced to integrate these heterogeneous kernel matrices in a more sophisticated manner. This approach enables the model to move beyond simple averaging or fixed weighting schemes, allowing it to adaptively modulate the contribution of each omics data type according to both the intrinsic characteristics of the data and its relevance to the learning task. Consequently, this enhances the expressiveness and robustness of the fused features. The specific fusion strategy is outlined as follows:

The linear embeddings of the three cell line omics (gene expression, CNA, mutation) are averaged to generated a unified shared fusion embedding, ***E***_***shared***_. This embedding captures the initial comprehensive features of a cell line across multi-omics dimensions and serves as the input for computing dynamic attention weights.

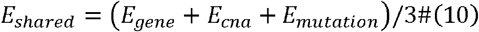

A compact neural network is utilized to learn dynamic attention weights for each cell line corresponding to the three omics data types. These weights are specific to each cell line and reflect the relative significance of each omics data type for individual cell lines within the given context.

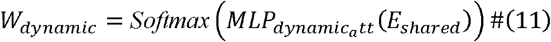

where ***MLP***_***dynamic_att***_ represents the linear layers and ReLU activation in the dynamic attention layer.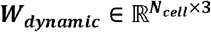, where each row ***w***_***dynamic***_**[*k***,:] corresponds to the dynamic contribution weights of the ***k* − *th*** cell line for the gene, CNA, and mutation kernels. The dynamic attention mechanism enables the model to ‘learn’ the distinct characteristics of each cell line. For instance, if a cancer cell line exhibits a highly aberrant gene expression pattern but relatively few mutations, the model can assign greater weight to the gene expression kernel via the learned attention. This personalized adjustment enhances the biological relevance of the fused similarity matrix and ensures it more accurately reflects the true similarity of each cell line. Moreover, when data are noisy or certain omics information is incomplete, the dynamic attention mechanism mitigates the impact of unreliable or redundant information, thereby enhancing the model’s robustness in handling complex and noisy biological data.

A set of learnable, globally shared parameter vectors ***w***_***static_param***_ ∈ ℝ^3^ (initialized as a 3-dimension tensor with all elements equal to one) is utilized. These parameters are then normalized using a softmax function to generate a fixed-proportion static weight vector ***w***_***static***_ ∈ ℝ^3^.

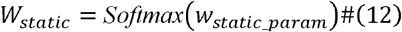

where ***w***_***static*[*i*]**_ represents the global static weight for the ***i* − *th*** kernel.

##### Mixed Weight Fusion

To balance the stability offered by static weights with the personalized adaptivity of dynamic weights, the final fusion weights are determined as a 1:1 combination of both approaches. This hybrid strategy not only captures local and subtle variations in the data but also preserves overall stability, thereby mitigating the risk of overfitting or information loss that may arise from relying solely on one weighting scheme. For the ***k* − *th*** cell line, the mixed weight for the ***i* − *th*** kernel, ***w***_***mixed*[*k***,***i*]**_, is computed as the sum of its static and dynamic weights, as follows:

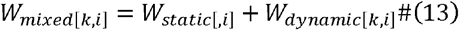

The ***k* − *th*** row of the final fused kernel matrix ***K***_***fused***_, which represents the similarity of the ***k* − *th*** cell line with all others, is obtained by elementwise multiplying each normalized kernel matrix ***K***_***norm***,***k***_ with its corresponding mixed weight ***w***_***mixed***,***k*,**_ followed by summing the resulting matrices:

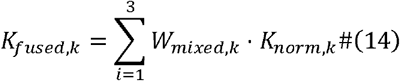

Through the aforementioned process, multi-modal fused features for cell lines and a Jaccard similarity matrix for drugs are generated. These features will then be utilized as inputs to construct and augment the heterogeneous graph module.

#### 2.2.3. Heterogeneous Graph Construction and Augmentation

This module represents the second core component of HGACL-DRP. As shown in **Fig. 3**, its primary function is to construct and refine the heterogeneous graph structure by leveraging the similarity information produced by the multi-modal feature fusion module. Additionally, it generates a structurally perturbed graph view for contrastive learning, providing a robust topological input for the subsequent graph convolutional network encoder. In biological networks, node connections frequently involve noisy or redundant information, which may adversely impact the performance of graph neural networks. Consequently, this module enhances the graph structure by incorporating a reliable neighbor filtering mechanism to maintain sparsity and ensure biological relevance. Finally, it integrates various types of connections into a unified heterogeneous adjacency matrix, on which structural perturbations are applied.

**Fig. 3.**
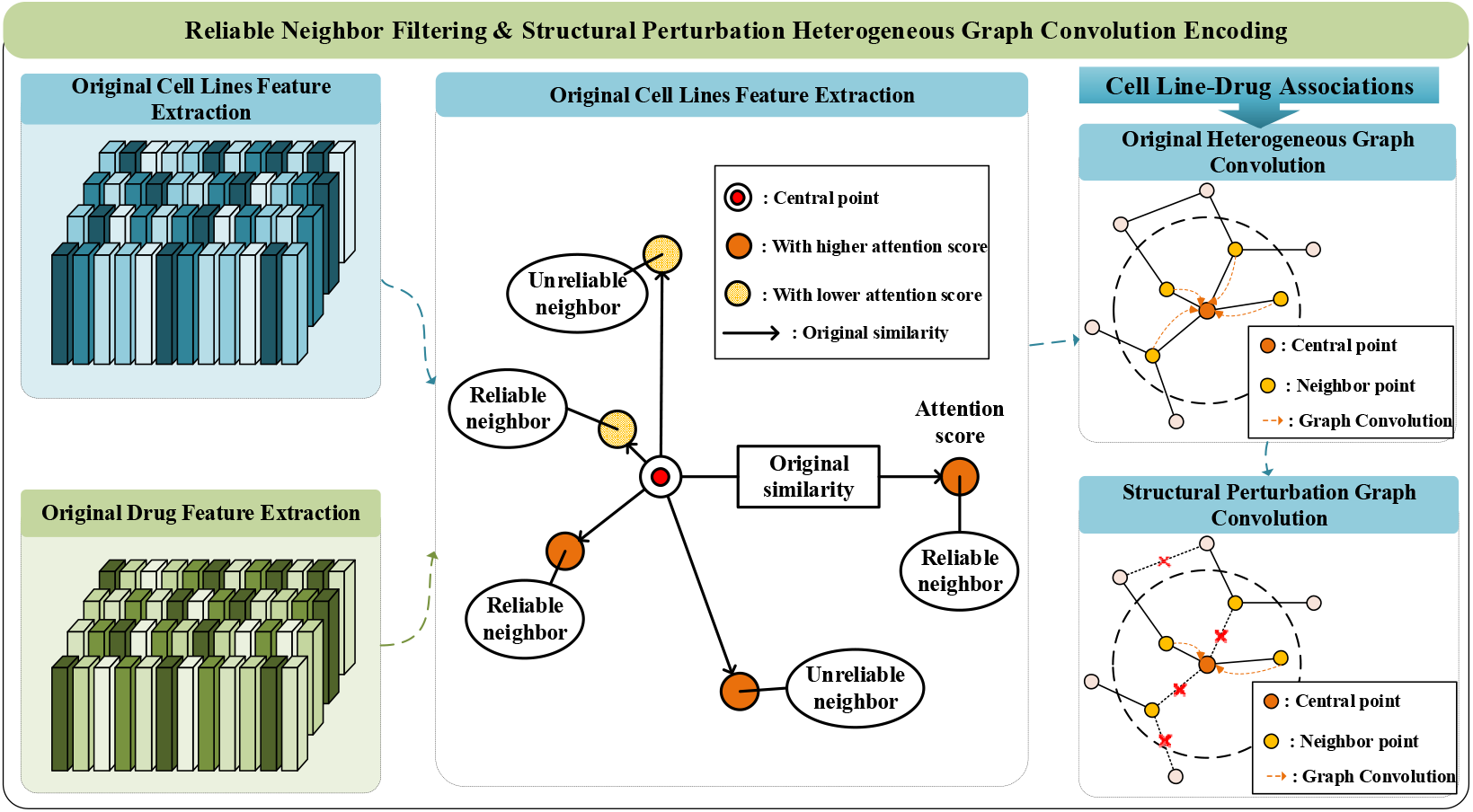
Heterogeneous Graph Construction and Augmentation.

##### Reliable Neighbor Filtering

To reduce noise in the similarity matrices and emphasize critical connections, we propose a novel reliable neighbor filtering mechanism based on node attention scores. This mechanism dynamically identifies and retains the most informative and “reliable” neighbor connections by considering both raw similarity scores and contextual relevance, rather than merely truncating based solely on raw similarity scores. Consequently, it constructs a more concise, efficient, and biologically interpretable graph structure, effectively minimizing interference from noise and redundant connections during information propagation. This ensures that selected neighbors are not only similar to the central node but also inherently information-rich entities, thereby enhancing the biological relevance of the graph topology. The mechanism is applied to both the fused cell line kernel matrix and the drug similarity matrix.

First, for each cell line and drug, we utilize an independent multi-head attention-based neighbor module to calculate their respective node attention scores. This module consists of a linear layer followed by a ReLU activation function, another linear layer, and finally a Sigmoid activation function:

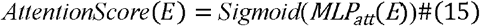

Where ***E*** represents the shared fusion embedding for cell lines or the data embedding for drugs. ***MLP***_***att***_ represents the linear layers and ReLU activation within the multi-head attention neighbor module. These attention scores reflect the relative importance or significance of each node within the feature space and are internally normalized through division by their sum.

For the relationship between a central node ***i*** and all its potential neighbors ***j***, we calculate a weighted similarity:

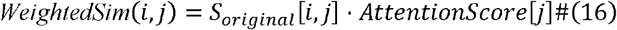

Where ***AttentionScore*[*j*]** is the attention score of the neighbor node ***j***, and ***S***_***original*[*I***,***j*]**_ is the original similarity between the central node ***i*** and its neighbor ***j***, derived from the fused cell line kernel matrix ***K***_***fused***_ or the normalized drug similarity kernel matrix ***K***_***drug***,***norm***_. The primary advantage of this weighting method lies in its ability to account for both the surface-level similarity between two nodes and the intrinsic importance of the neighbor node. This ensures that nodes designated as ‘reliable neighbors’ are not only similar to the central node but also exhibit high levels of informativeness or credibility independently.

Subsequently, for each central node ***i***, we identify the indices of the top-k (with k set to10 in the experiments) neighbors that exhibit the highest weighted similarity scores among all its neighbors:

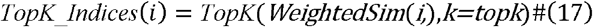

Only the values in the original similarity matrix corresponding to the neighbors selected in ***TopK*_*Indices*** are retained; all other positions are set to zero. This process is applied independently to the cell line and drug similarity matrices.

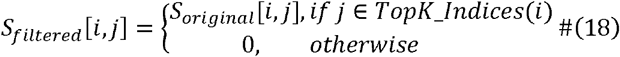

##### Heterogeneous Adjacency Matrix Construction and Structural Perturbation

This stage is designed to construct the model’s initial heterogeneous graph input and generate a structurally perturbed graph view for contrastive learning. Specifically, the filtered cell line and drug similarity matrices, together with the original binarized cell line-drug associations, are integrated into a unified heterogeneous adjacency matrix 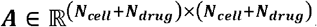.The constructed matrix ***A*** is subsequently symmetrically normalized to ensure the numerical stability and effectiveness of the graph convolution operations, yielding the normalized adjacency matrix 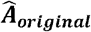 for encoding the original graph view:

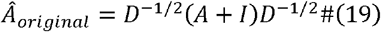

where ***D*** is the degree matrix of ***A*+*I***, and ***I*** is the identity matrix.

We generate the structurally perturbed graph by applying random edge dropping to the original cell line-drug associations. Specifically, a fraction ***α*** (referred to as the structural perturbation coefficient) of the existing edges (i.e., elements with a value of 1 in the original cell line-drug association matrix) are randomly selected and set to 0. This process yields a perturbed cell line-drug association matrix. Subsequently, this perturbed matrix is combined with the precomputed cell line and drug similarity matrices to reconstruct a structurally perturbed heterogeneous adjacency matrix. Finally, the newly reconstructed matrix undergoes symmetric normalization to yield the normalized adjacency matrix 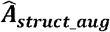, which encodes the structural perturbation graph view.

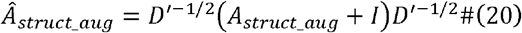

Where ***A***_***struct*_*aug***_ is the heterogeneous graph constructed based on the perturbed cell line-drug association matrix. This structural perturbation is advantageous as it simulates potential missing links or noise in biological networks, thus compelling the graph encoder to learn node representations that are more resilient to changes in graph topology. Consequently, this approach improves the model’s generalization capability.

Through the aforementioned steps, we derive the normalized heterogeneous adjacency matrices for different views: 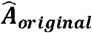 corresponding to the original view and 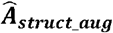 associated with the structural perturbation view. Subsequently, these matrices, together with the fused node features, are fed as input to the graph encoder (GEncoder) to generate the respective node embeddings:

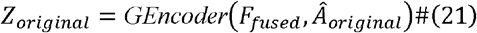

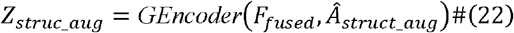

where ***F***_***fused***_ represents the fused cell line and drug node features obtained through a linear transformation. The GEncoder functions as a heterogeneous graph convolutional operator, specifically designed to handle the heterogeneous graph structure we constructed earlier. Its calculation process strictly adheres to the general information aggregation framework of graph neural networks and leverages the normalized heterogeneous adjacency matrix that was generated in the preceding section.

For any node—cell line or drug—GEncoder aggregates features from all neighbors. Exploiting the three edge types (cell-cell, drug-drug, cell-drug) encoded in the heterogeneous adjacency matrix, it updates each representation with both homogeneous and heterogeneous context. This simultaneous integration captures intricate network interactions and prevents the graph from being mistaken for a pseudo-homogeneous structure.

#### 2.2.4. Dual-Perturbation Contrastive Self-Supervised Learning

As shown in Fig. 4, the primary goal of this module is to learn more robust and discriminative node embeddings for cell lines and drugs via a Dual-Perturbation contrastive self-supervised learning framework. This objective becomes especially critical when addressing the challenges posed by sparse drug response data and limited labeled samples. In contrast to traditional supervised learning, which relies heavily on large-scale annotated datasets, contrastive learning leverages self-supervised tasks to extract intrinsic information directly from the data, thereby improving feature representation capabilities in scenarios with scarce or no labels. We therefore construct three distinct views—an original view, a structural perturbation view, and a feature perturbation view—all specifically tailored for DRP tasks. These views are then integrated through a tailored InfoNCE (Information Noise-Contrastive Estimation) loss function, which maximizes the consistency between semantically similar samples across different views while effectively distinguishing dissimilar ones.

**Fig. 4.**
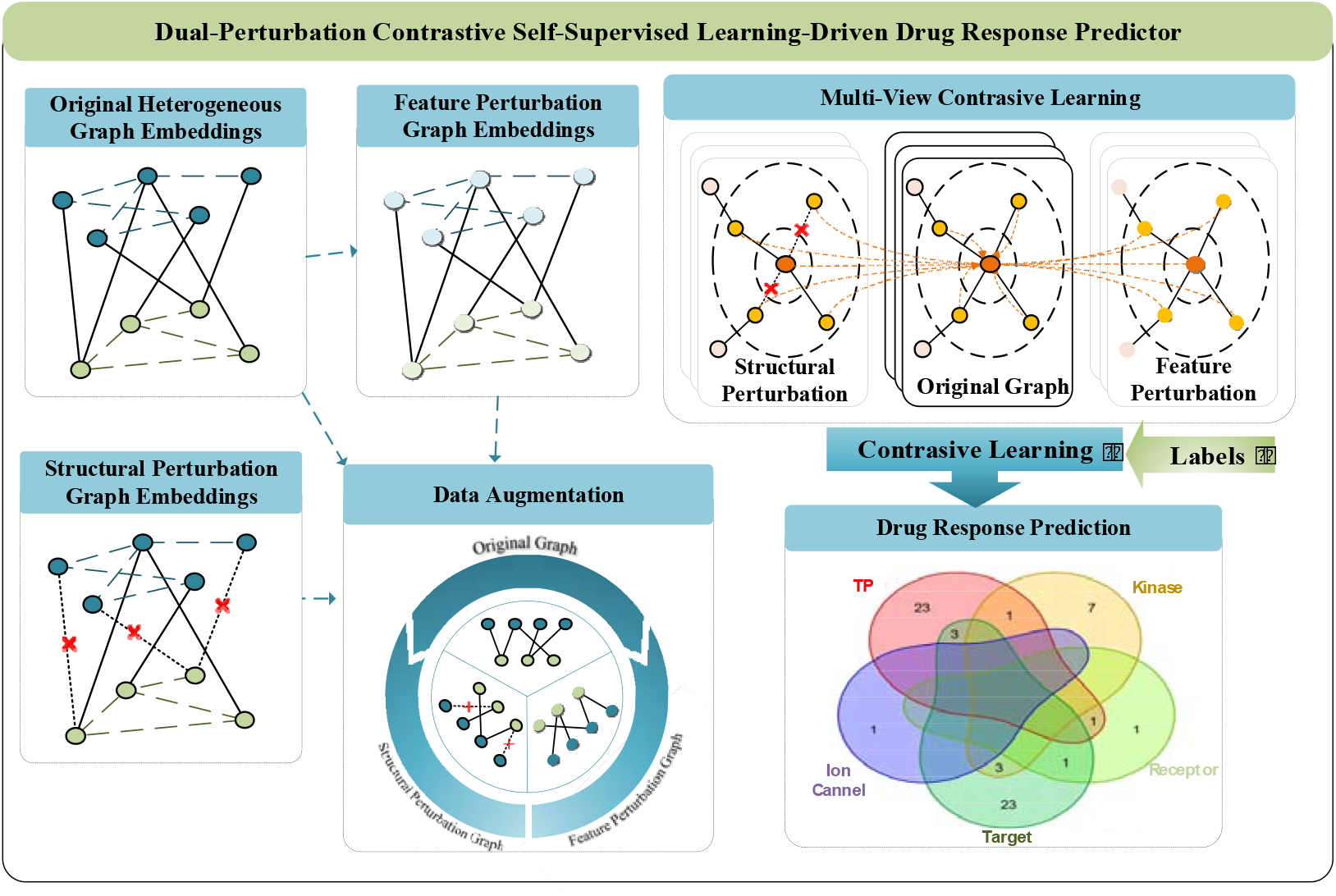
Dual-Perturbation Contrastive Self-Supervised Learning.

##### Feature Perturbation and Data Augmentation

To enhance robustness against biological noise and network incompleteness, we simultaneously introduce feature-level Gaussian noise and structural random edge dropout into our contrastive learning framework. This dual-perturbation strategy has been empirically shown to substantially improve representation generalization in large-scale graph contrastive studies: GraphCL [34] systematically compared four types of graph perturbations and observed consistent and significant performance gains from additive Gaussian noise and random edge removal across seven public datasets; GCA [35] further adapted these two perturbations to different nodes, achieving an average AUC improvement of 3.2% on heterogeneous graphs such as biological networks. Motivated by these findings, we inject Gaussian noise into node embeddings to mimic omics measurement errors and randomly drop cell–drug interaction edges with a learned probability to simulate network sparsity, thereby forcing the encoder to learn stable representations that are insensitive to minor variations.

As detailed in Section 2.2.3, the GEncoder has already generated node embeddings for two graph views: the original graph view ***Z***_***original***_ and the structural perturbation graph view ***Z***_***struct_augZ***_ based on distinct graph structures. Expanding on this foundation, this module introduces a third view—the feature perturbation view***Z***_***feat_aug***_—to strengthen the robustness of the node embeddings from multiple angles. This view is constructed by superimposing Gaussian noise directly onto to the node embeddings produced by the original graph encoder ***Z***_***original***_. The significance of this perturbation lies in its ability to emulate potential minor measurement errors or biological variability in node features, thereby driving the model to learn representations that are robust to feature-level noise. This approach not only enhances the robustness of the learned features but also improves the model’s adaptability to real-world data variations:

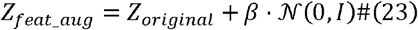

where ***β*** denotes the noise intensity coefficient. ***N* (0**,***I*)**represents a random tensor sampled from a standard normal distribution, with the same shape as ***Z***_***original***_. This ensures the randomness and diversity of the perturbation introduced into the system.

This study incorporates contrastive learning to enhance the model’s capacity for recognizing complex drug response patterns and improving its generalization ability. The essence of this method lies in defining and comparing sample pairs across multiple views, enabling the model to automatically learn robust and meaningful feature representations from the data. In DRP task, it is critical to accurately define positive, negative, and “neutral” pairs for each node. This ensures that the model learns information more aligned with underlying biological patterns—specifically, identifying which relationships should be “pulled closer” (similar), which should be “pushed apart” (dissimilar), and which should remain unoptimized.

##### Positive Pairs

These pairs represent relationships that exist in the initially known associations (including self-loops or established cell-drug sensitivity links). For these pairs, the model aims to bring their embeddings closer together in the latent space. This approach facilitates the model’s ability to identify and reinforce similarities influenced by multiple factors, which is essential for improving generalization, especially when handling sparse or noisy data. For example, if cell line A exhibits sensitivity to drug B, their respective representations across different views form a positive pair, prompting the model to learn how to align their embeddings more effectively.

##### “Neither-Positive-Nor-Negative” Pairs

This type of pair represents a critical innovation in the model, as it integrates biological prior knowledge into the contrastive learning process. Specifically, these pairs are identified through the “reliable neighbor filtering mechanism.” For such pairs, the model neither enforces their embeddings to be closer nor drives them further apart. The advantage of this strategy lies in its ability to prevent over-optimization of relationships that are already explicit, strongly correlated, or well-established from a biological standpoint. Consequently, the model can allocate its limited optimization resources and gradients more effectively toward uncovering and reinforcing “non-obvious” associations via the self-supervised task, thus enhancing overall learning efficiency and generalization.

##### Negative Pairs

These pairs correspond to connections that are neither part of any established cell-drug sensitivity associations nor identified as “reliable.” For these pairs, the model seeks to increase the distance between their embeddings in the latent space. This enables the model to effectively differentiate between unrelated entities, thereby enhancing its discriminative capability and avoiding the misclassification of samples that should not be deemed similar.

The InfoNCE loss function, expressed as a log-likelihood ratio, quantifies the similarity of an anchor with its positive sample relative to its similarity with all other samples (including positive and negative ones). For an anchor ***i***, with a set of positive samples ***P*(*i*)** and a set of negative samples ***N*(*i*)** in another contrastive view, its contribution to the contrastive loss is derived as follows:

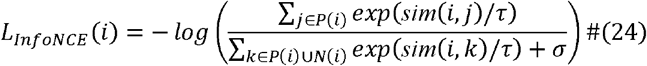

where **τ** is the temperature parameter and ***σ*** is a small constant added to ensure numerical stability. By minimizing this loss, the model is incentivized to bring the embeddings of positive pairs closer while simultaneously pushing those of negative pairs further apart. We compute the InfoNCE loss for three distinct contrastive tasks, thereby comprehensively enhancing the quality of the embeddings from multiple perspectives:

- ***L***_***feat***_: Measures the consistency between the original view ***Z***_***original***_ and the feature perturbation view ***Z***_***feat_aug***_.
- ***L***_***struct***_: Measures the consistency between the original view ***Z***_***original***_ and the structural perturbation view ***Z***_***struct_aug***_.
- ***L***_***orig_intra***_: Measures the internal consistency within the original view ***Z***_***original***_ by contrasting nodes with their own neighbors in the graph.

The total contrastive loss, ***L***_***contrastive***_, is the average of these three components.

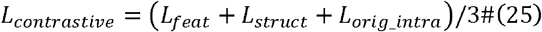

This multi-task contrastive objective ensures that the learned embeddings are simultaneously robust to feature-level noise, topological changes, and capable of preserving local neighborhood information. The primary strength of this Dual-Perturbation contrastive learning strategy is its capacity to drive the model to learn node representations that are highly resilient to biological data variations across different perturbation granularities (both feature-level and structural-level). Additionally, the refined definition of positive and negative samples enhances the model’s discriminative power for deciphering the complex mechanisms underlying drug response. This self-supervised learning paradigm markedly enhances the model’s generalization capability even with limited labeled data, enabling it to uncover deep, implicit patterns within the data and extract richer, more discriminative features for drug response prediction.

#### 2.2.5. Drug Response Prediction

This module leverages the robust and discriminative cell line and drug node embeddings learned via the Dual-Perturbation contrastive self-supervised learning module (Section 2.2.4) to accurately predict the sensitivity of cell lines to drugs. The prediction score for a cell line-drug response is calculated by evaluating the Pearson Correlation Coefficient between their transformed embeddings. This correlation coefficient, constrained within the range [-1, 1], is subsequently passed through a Sigmoid activation function to yield in a probability value within [0, 1]. This value quantitatively reflects the sensitivity of the cell line to the drug.

In this study, contrastive loss and supervised loss are integrated within an end-to-end framework for learning feature representations. The contribution of the supervised task to the overall loss is quantified by the cross-entropy loss function, which can be formulated as follows:

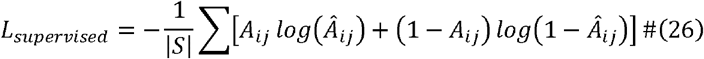

where ***S*** represents the training set of the same batch, ***A***_***ij***_ the actual label values between nodes, and 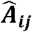 predicted values between nodes.

To simultaneously achieve drug resistance prediction and contrastive learning, the losses from contrastive learning and supervised learning are integrated, optimizing the following loss function:

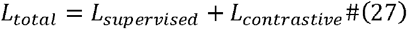

## 3. Experimental Results and Discussion

In this section, a thorough evaluation of the HGACL-DRP model’s performance is conducted via a series of comprehensive experiments. The comparison algorithms and the metrics employed for quantitative assessment are first described. An in-depth analysis of HGACL-DRP’s predictive capabilities relative to state-of-the-art methods follows. Additionally, ablation studies are carried out to elucidate the contributions of each module within the proposed framework and hyperparameter sensitivity analyses are performed to highlight the robustness of the model. Lastly, a biological interpretation of the findings is offered through drug analysis and visualization of learned embeddings, thereby revealing the model’s clinical relevance and discriminative power.

### 3.1. Comparison Algorithms and Evaluation Metrics

To comprehensively evaluate our proposed HGACL-DRP model, we selected seven state-of-the-art drug-response-prediction algorithms for comparison. These algorithms adopt distinct methodologies for data fusion, network construction, and learning paradigms, as detailed below.

#### DeepDRK [18] (2021)

A deep learning framework based on kernel functions for multi-omics data fusion. It integrates heterogeneous information by constructing kernel similarity matrices from multi-source data and utilizes a deep neural network for prediction.

#### MOFGCN [9] (2022)

This method calculates cell line similarity by fusing multi-omics data, combines it with drug similarity to construct a heterogeneous network, and finally learns latent features via a graph convolutional network to predict drug response.

#### GraphCDR [14] (2022)

It combines graph neural networks with contrastive learning, enhancing the model’s generalization ability by contrasting the embeddings of a “sensitivity graph” and its opposing “resistance graph”.

#### NIHGCN [10] (2022)

This model adopted a parallel heterogeneous graph convolutional network based on Neighborhood Interaction (NI), designed to simultaneously aggregate node-level and element-level neighbor features to capture more detailed interaction information.

#### TSGCNN [11] (2023)

It innovatively performs graph convolution first within homogeneous cell line and drug feature spaces to diffuse similarity, and then aggregates information on the heterogeneous network.

#### MultiDRP [12] (2024)

It constructs a hierarchical attention network that captures external relationships between entities via a graph attention network and reinforces internal correlations among features using a multi-head self-attention network, thereby achieving the fusion of multi-scale relationships.

#### ASGCL [15] (2025)

This model introduces an adaptive sparse mapping-based graph contrastive learning network. Its core is a GraphMorpher module that performs graph augmentation through node attribute masking and topological pruning, and it designs multi-level contrastive learning tasks to enhance feature discrimination.

To guarantee reproducibility and fairness, we implemented all algorithms within the same framework. Specifically, we adopted five-fold cross-validation for every model. Importantly, we executed all comparison algorithms—together with our HGACL-DRP—on the identical, pre-processed input data under this unified protocol, rather than citing performance figures from their original papers. This standardized design yields a direct and unbiased assessment of predictive performance.

To quantitatively and multi-dimensionally assess the predictive performance of HGACL-DRP and all comparison algorithms, six widely recognized evaluation metrics for binary classification tasks were utilized. Specifically, the Area Under the Receiver Operating Characteristic curve (AUC) provides a comprehensive measure of the model’s ability to distinguish between positive and negative samples across all thresholds, with values closer to 1 indicating superior discriminative performance. Accuracy (ACC) reflects the proportion of correctly classified samples (both sensitive and resistant) relative to the total number of samples at a given threshold. Furthermore, Precision quantifies the proportion of truly sensitive samples among those predicted as sensitive; Recall measures the proportion of correctly identified sensitive samples among all actual sensitive samples; and the F1-Score, as the harmonic mean of Precision and Recall, offers a balanced evaluation of the model’s precision and recall capabilities. Finally, the Matthews Correlation Coefficient (MCC), a robust metric that remains effective even in cases of imbalanced class distributions, was utilized, where higher values indicate better prediction performance.

### 3.2 Performance Comparison

To evaluate HGACL-DRP’s effectiveness, a performance comparison was conducted against seven state-of-the-art baseline methods using the CCLE and GDSC datasets, ensuring identical input features and data preprocessing. HGACL-DRP consistently outperformed all competing methods across key evaluation metrics on both datasets.

On the CCLE dataset, HGACL-DRP achieved an AUC of 0.9548, surpassing all comparison models, including ASGCL (AUC of 0.8828). This indicates HGACL-DRP’s superior ability to distinguish between sensitive and resistant samples. HGACL-DRP also showed superiority in F1-Score (0.9194) and MCC (0.8368), outperforming ASGCL (F1-Score: 0.8268) and NIHGCN (F1-Score: 0.7954). Compared to graph convolutional models without contrastive learning (e.g., MOFGCN AUC 0.8624, TSGCNN AUC 0.8674) and earlier deep learning models (e.g., DeepDRK AUC 0.7985), HGACL-DRP demonstrated comprehensive performance improvements, validating its innovative graph structure optimization and Dual-Perturbation contrastive learning framework.

On the larger and more challenging GDSC dataset, HGACL-DRP maintained its leading position with an exceptional AUC of 0.9899, exceeding ASGCL’s AUC of 0.9762. It also achieved F1-Score of 0.9708 and MCC of 0.9414, surpassing ASGCL’s scores of 0.9523 (F1) and 0.9038 (MCC). HGACL-DRP showed a substantial performance advantage over MultiDRP, whose F1-Score and MCC on GDSC were 0.4887 and 0.4330, respectively. This indicates that HGACL-DRP’s “graph optimization + contrastive learning” framework is more effective than MultiDRP’s pure attention mechanism for drug response prediction. Overall, HGACL-DRP achieved the best numerical performance across both datasets, validating its design methodology and setting a new benchmark for precision drug response prediction.

To intuitively demonstrate the performance advantages of HGACL-DRP over other baseline methods, the Receiver Operating Characteristic (ROC) curves and Precision-Recall (PR) curves for all models on the CCLE and GDSC datasets are presented in **Fig. 5**. From the ROC curves, it is evident that the curve representing HGACL-DRP (solid blue line) consistently outperforms the curves of all other comparison methods across both datasets by occupying the top-left corner. This indicates that for any given False Positive Rate, HGACL-DRP achieves the highest True Positive Rate, thereby showcasing the strongest discriminative power between sensitive and resistant samples. Furthermore, the AUC values listed in the legend further support this conclusion, with HGACL-DRP attaining an AUC of 0.955 on CCLE and 0.990 on GDSC, which are substantially higher than those of all competing algorithms. The PR curve analysis provides compelling evidence for the model’s performance in real-world scenarios with class imbalance, where resistant samples significantly outnumber sensitive ones. HGACL-DRP’s curve consistently shows dominance, indicating its ability to achieve exceptionally high prediction precision while recalling a larger proportion of sensitive samples. The model’s outstanding Area Under the PR Curve (AUPR) values of 0.948 on CCLE and 0.981 on GDSC further substantiate the high reliability of drug-cell line pairs predicted as “sensitive” by HGACL-DRP, which is critically important for guiding experimental validation and clinical applications.

**Fig. 5.**
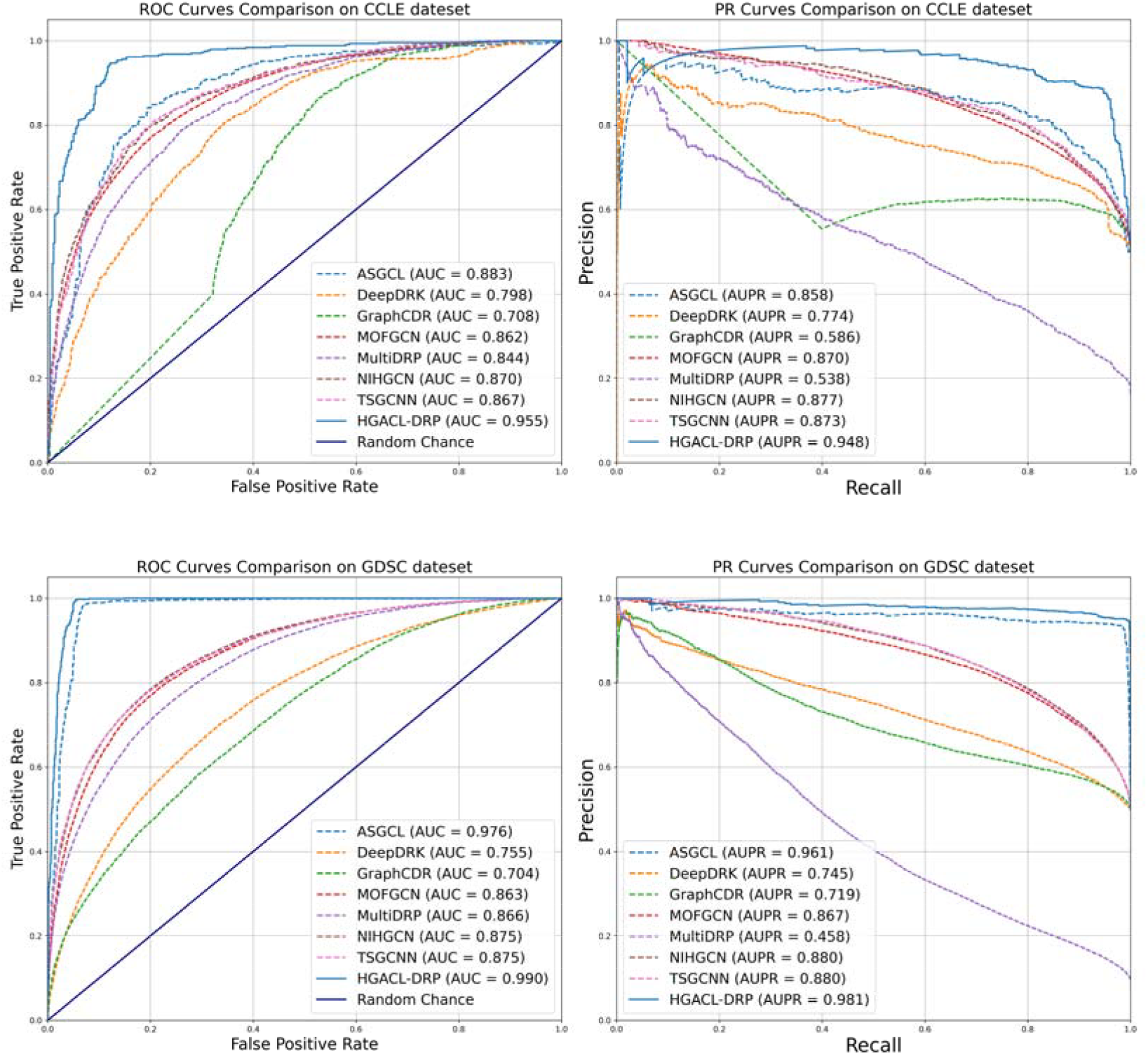
ROC and PR curves of HGACL and its comparative algorithms on CCLE and GDSC datasets.

### 3.3 Ablation Experiments

To intuitively evaluate the contribution of each model component, we generated a radar chart based on the ablation study results, as shown in **Fig. 6**. In both the CCLE and GDSC datasets, it is clear that the polygon corresponding to the complete HGACL-DRP model (solid blue line) exhibits the largest area and completely encloses all other polygons representing the model variants.

**Fig. 6.**
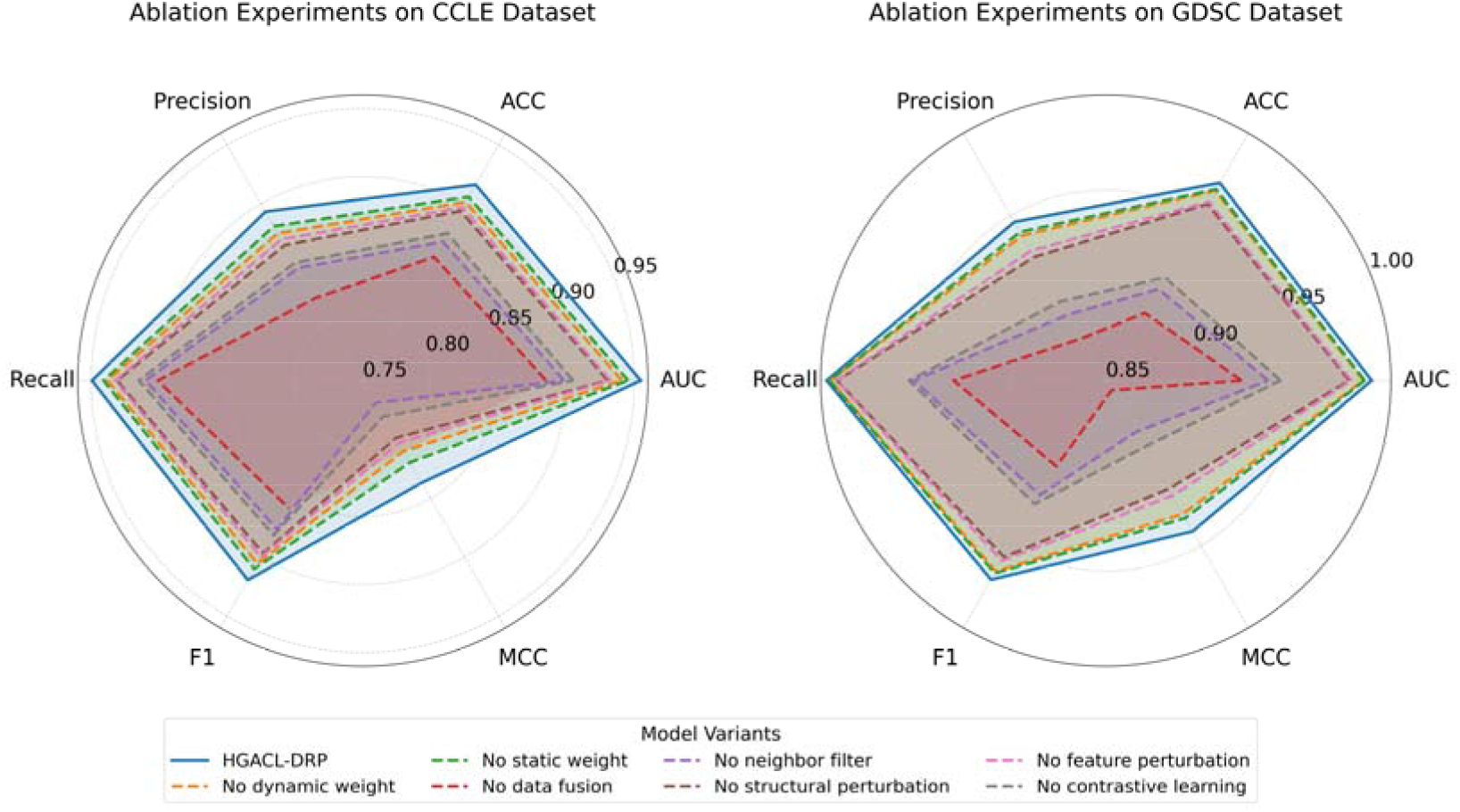
Radar charts of ablation experiments on CCLE and GDSC datasets.

This visual envelopment strongly substantiates the consistent superiority of the complete model across all six evaluation metrics (ACC, AUC, MCC, F1, Recall, Precision). For each variant with a specific component removed, the corresponding polygon contracts toward the center, accompanied by a reduction in area, which intuitively illustrates that the removal of any single module leads to a decline in the model’s overall performance.

Notably, the removal of the ‘data fusion’ (No data fusion), ‘reliable neighbor filtering’ (No neighbor filter), and ‘contrastive learning’ (No contrastive learning) modules s leads to the most significant reduction in the size of the corresponding polygons(dashed red, purple, and grey lines). This visually underscores the critical role these components play in enabling the model’s superior performance. Consequently, the radar chart provides clear and convincing visual evidence for the robust design of the HGACL-DRP model and the efficacy of its individual components.

### 3.4 Hyperparameter Evaluation

We conducted a sensitivity analysis on the structural perturbation coefficient () and feature perturbation coefficient () to understand their impact on model performance. As shown in **Fig. 7**, both parameters exhibit an inverted U-shaped performance curve, indicating an optimal balance point for data augmentation in self-supervised learning. Initially, increasing perturbation improves performance, as moderate structural perturbation helps the model capture robust topological patterns, and feature perturbation enhances resilience to noise. However, beyond optimal thresholds (=0.2, =0.1), performance declines because excessive perturbation leads to information loss. This trend demonstrates that optimal performance is achieved by balancing robustness enhancement and information integrity preservation.

**Fig. 7.**
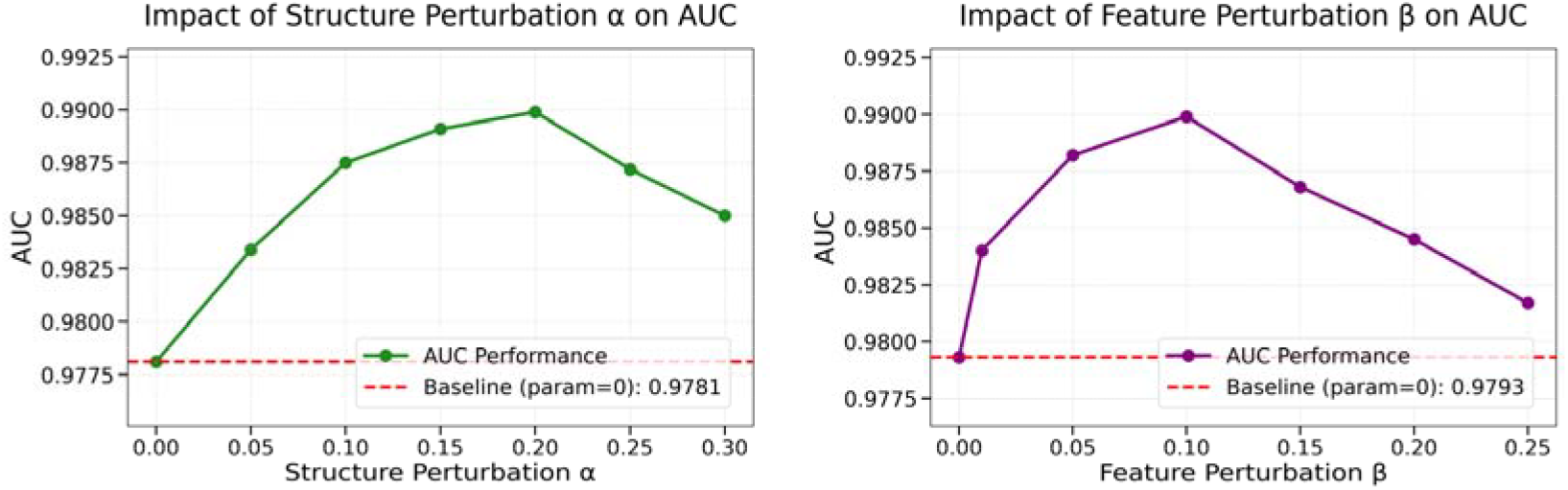
Hyperparameter Evaluation on CCLE and GDSC datasets.

**Fig. 8.**
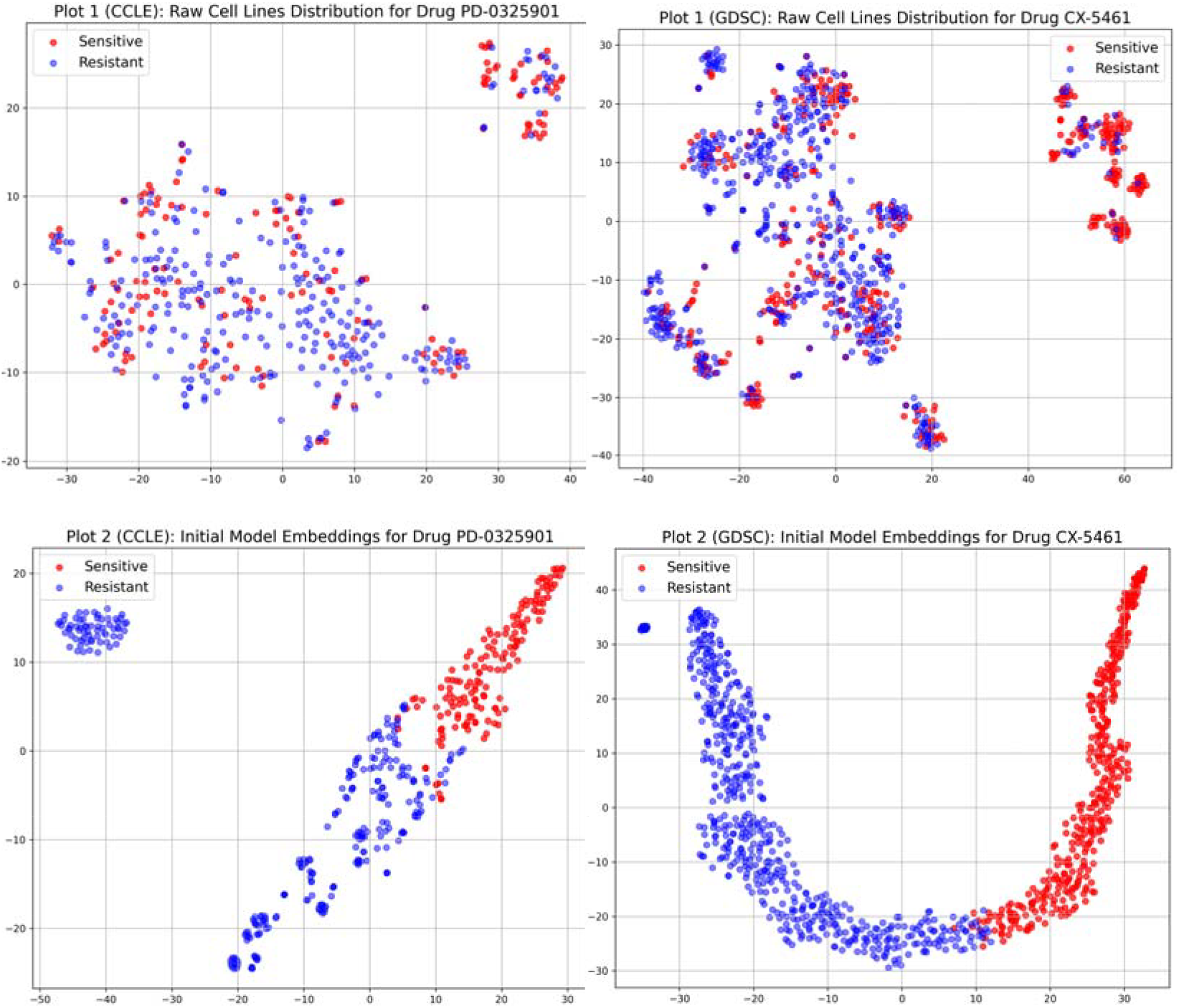
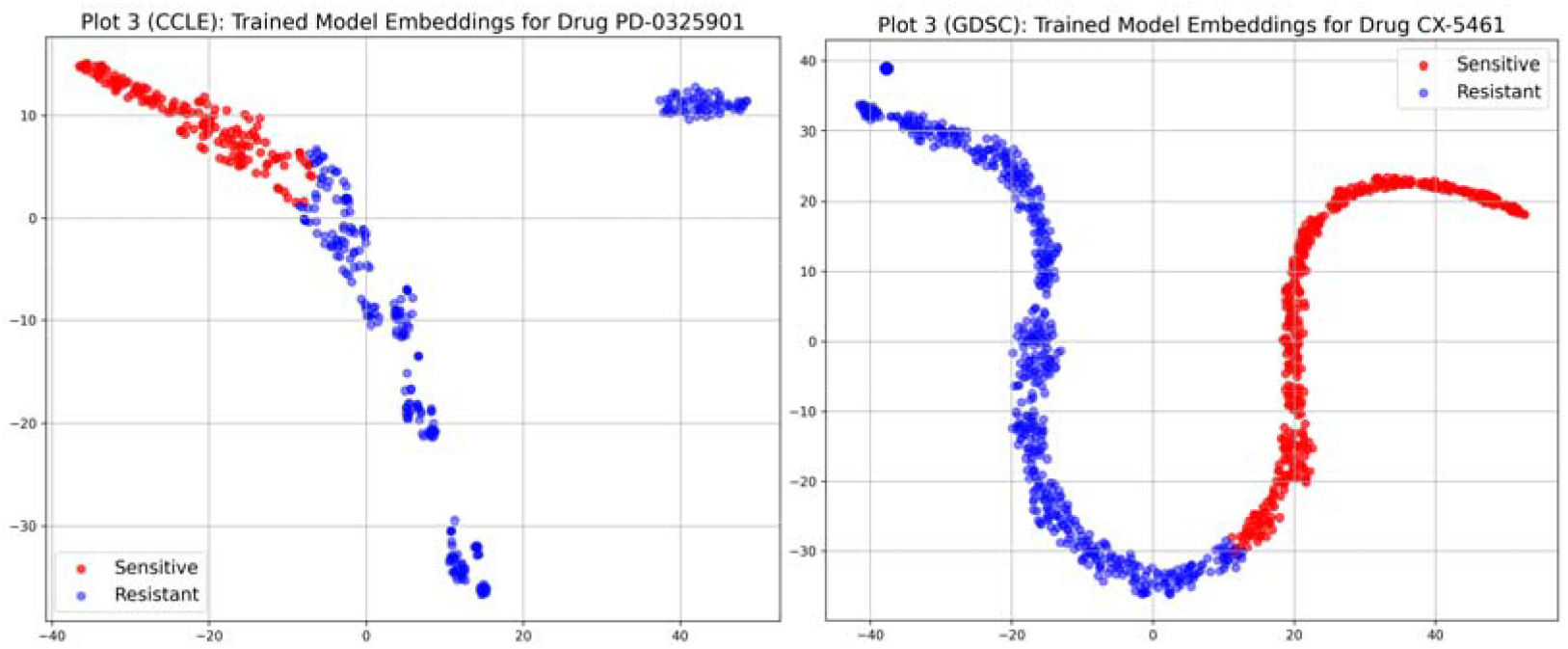
Distribution of cell line pairs based on different training stages of drug network.

### 3.5 Drug Analysis

To further validate the practical application value of the HGACL-DRP model and the biological significance of its predictions, we conducted an in-depth analysis of the top 10 drugs predicted to have the highest number of sensitive cell lines in both the CCLE and GDSC databases. Specific details regarding these drugs are summarized in **Table 3**. Our analysis revealed that the majority of the drugs identified by the model are potent anti-cancer agents, either clinically approved or currently under clinical investigation. Their targets encompass a broad spectrum of core cancer signaling pathways, including well-established kinase inhibitors as well as cutting-edge regulators of apoptosis and epigenetic control.

**Table 1.**
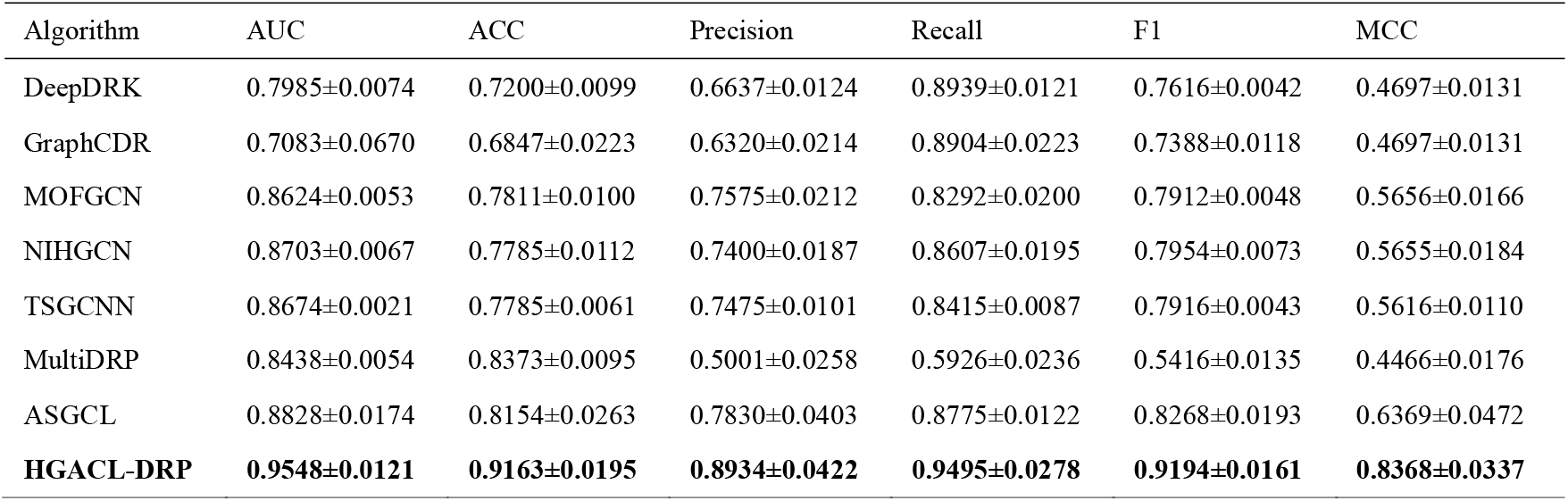
Comparison of HGACL-DRP and other classification models on CCLE datasets.

| Algorithm | AUC | ACC | Precision | Recall | F1 | MCC |
| --- | --- | --- | --- | --- | --- | --- |
| DeepDRK | 0.7985±0.0074 | 0.7200±0.0099 | 0.6637±0.0124 | 0.8939±0.0121 | 0.7616±0.0042 | 0.4697±0.0131 |
| GraphCDR | 0.7083±0.0670 | 0.6847±0.0223 | 0.6320±0.0214 | 0.8904±0.0223 | 0.7388±0.0118 | 0.4697±0.0131 |
| MOFGCN | 0.8624±0.0053 | 0.7811±0.0100 | 0.7575±0.0212 | 0.8292±0.0200 | 0.7912±0.0048 | 0.5656±0.0166 |
| NIHGCN | 0.8703±0.0067 | 0.7785±0.0112 | 0.7400±0.0187 | 0.8607±0.0195 | 0.7954±0.0073 | 0.5655±0.0184 |
| TSGCNN | 0.8674±0.0021 | 0.7785±0.0061 | 0.7475±0.0101 | 0.8415±0.0087 | 0.7916±0.0043 | 0.5616±0.0110 |
| MultiDRP | 0.8438±0.0054 | 0.8373±0.0095 | 0.5001±0.0258 | 0.5926±0.0236 | 0.5416±0.0135 | 0.4466±0.0176 |
| ASGCL | 0.8828±0.0174 | 0.8154±0.0263 | 0.7830±0.0403 | 0.8775±0.0122 | 0.8268±0.0193 | 0.6369±0.0472 |
| **HGACL-DRP** | **0.9548±0.0121** | **0.9163±0.0195** | **0.8934±0.0422** | **0.9495±0.0278** | **0.9194±0.0161** | **0.8368±0.0337** |

**Table 2.** Comparison of HGACL-DRP and other classification models on GDSC datasets.

| Algorithm | AUC | ACC | Precision | Recall | F1 | MCC |
| --- | --- | --- | --- | --- | --- | --- |
| DeepDRK | 0.7554±0.0021 | 0.6611±0.0036 | 0.6166±0.0041 | 0.8522±0.0064 | 0.7155±0.0016 | 0.3487±0.0055 |
| GraphCDR | 0.7039±0.0366 | 0.6118±0.0257 | 0.5718±0.0201 | 0.9012±0.0149 | 0.6992±0.0110 | 0.2731±0.0444 |
| MOFGCN | 0.8627±0.0016 | 0.7741±0.0032 | 0.7469±0.0089 | 0.8295±0.0119 | 0.7859±0.0020 | 0.5517±0.0053 |
| NIHGCN | 0.8752±0.0008 | 0.7889±0.0015 | 0.7715±0.0025 | 0.8207±0.0018 | 0.7954±0.0012 | 0.5789±0.0029 |
| TSGCNN | 0.8754±0.0006 | 0.7897±0.0007 | 0.7733±0.0032 | 0.8198±0.0046 | 0.7959±0.0006 | 0.5805±0.0011 |
| MultiDRP | 0.8662±0.0006 | 0.8983±0.0024 | 0.4686±0.0111 | 0.5112±0.0121 | 0.4887±0.0032 | 0.4330±0.0037 |
| ASGCL | 0.9762±0.0043 | 0.9517±0.0076 | 0.9399±0.0084 | 0.9651±0.0131 | 0.9523±0.0077 | 0.9038±0.0154 |
| **HGACL-DRP** | **0.9899±0.0055** | **0.9699±0.0102** | **0.9462±0.0201** | **0.9969±0.0026** | **0.9708±0.0097** | **0.9414±0.0195** |

**Table 3.** Drug sensitivity analysis.

| Rank | CCLE |  |  | GDSC |  |  |
| --- | --- | --- | --- | --- | --- | --- |
|  | Drug | CID | Count | Drug | CID | Count |
| 1 | PD-0325901 | 9826528 | 155 | CX-5461 | 25257557 | 502 |
| 2 | RAF265 | 11656518 | 133 | Y-39983 | 11507964 | 327 |
| 3 | TAE684 | 16038120 | 125 | QL-XI-92 | 73265214 | 303 |
| 4 | ZD-6474 | 3081361 | 115 | GSK690693 | 16725726 | 277 |
| 5 | TKI258 | 135611162 | 115 | AZD6244 | 10127622 | 274 |
| 6 | AEW541 | 11476171 | 113 | Tubastatin A | 49850262 | 268 |
| 7 | PD-0332991 | 5330286 | 105 | Trametinib | 11707110 | 249 |
| 8 | AZD6244 | 10127622 | 104 | ABT-263 | 24978538 | 174 |
| 9 | Nilotinib | 644241 | 102 | Nutlin-3a | 11433190 | 159 |
| 10 | AZD0530 | 10302451 | 99 | A-770041 | 9549184 | 133 |

Among the drugs identified by the HGACL-DRP model, a significant proportion are targeted therapeutic agents that are already well-established in clinical practice. For instance, Trametinib [21] and Selumetinib (AZD6244) [22] are both highly effective MEK inhibitors. Trametinib is approved for treating BRAF-mutant melanoma, whereas Selumetinib is indicated for neurofibromatosis type 1. Palbociclib (PD-0332991), a CDK4/6 inhibitor, has demonstrated remarkable efficacy in the treatment of hormone receptor-positive breast cancer [23]. Nilotinib, a second-generation BCR-ABL tyrosine kinase inhibitor, serves as a first-line therapy for chronic myeloid leukemia (CML) [24]. Furthermore, Vandetanib (ZD-6474), a multi-targeted tyrosine kinase inhibitor, is approved for the treatment of medullary thyroid cancer [25]. The capacity of HGACL-DRP to precisely identify these clinically validated drugs with proven therapeutic efficacy highlights the model’s accuracy and its relevance to clinical applications.

In addition to established drugs, the model successfully identified a panel of promising candidate drugs currently under clinical investigation. For instance, RAF265 [26], a dual inhibitor of RAF and VEGFR2, and Dovitinib (TKI258) [27], a multi-targeted inhibitor of FGFR and VEGFR, are both being evaluated in ongoing clinical trials for various solid tumors. Saracatinib (AZD0530), an inhibitor of the Src family kinases, is also actively being investigated for its therapeutic potential across multiple cancer types [28]. The incorporation of these agents demonstrates that HGACL-DRP not only recognizes known effective therapies but also uncovers cutting-edge anti-cancer strategies at the forefront of research.

More notably, our model successfully identified several cutting-edge compounds acting on innovative targets, thereby underscoring its potential to uncover novel anti-cancer mechanisms. For example, CX-5461 is a newly developed RNA polymerase I (Pol I) inhibitor that induces cancer cell apoptosis by suppressing ribosome biogenesis and has demonstrated robust preclinical efficacy in hematological malignancies [29]. Both Nutlin-3a [30] and ABT-263 (Navitoclax) [31] are pivotal molecules targeting the apoptosis pathway: the former reactivates the p53 pathway by inhibiting the MDM2-p53 interaction, while the latter directly triggers apoptosis by inhibiting Bcl-2 family proteins. Additionally, Tubastatin A, a highly selective HDAC6 inhibitor, represents a promising direction in anti-cancer therapy via epigenetic regulation [32]. GSK690693, as a pan-Akt kinase inhibitor, targets the frequently dysregulated PI3K/Akt survival pathway in tumors [33]. The model’s capacity to identify these compounds with diverse mechanisms of action highlights its ability to recognize a broader spectrum of cancer cell vulnerabilities beyond conventional kinase inhibition.

In summary, HGACL-DRP successfully identified a series of anti-cancer drugs with high clinical relevance and diverse mechanisms of action. These include not only multiple marketed drugs that have demonstrated efficacy but also numerous promising candidates currently under clinical investigation, as well as cutting-edge compounds targeting innovative pathways. These findings robustly validate HGACL-DRP as a powerful and reliable tool for drug response prediction, highlighting its significant potential in screening effective drugs, accelerating novel drug discovery, and guiding personalized precision medicine.

### 3.6 Visualization Analysis

To visualize the discriminative capability of features learned by the HGACL-DRP model, t-SNE was used for 2D visualization of cell line embeddings for specific drugs. The results show a transformation from “chaos” to “order”. In the original feature space (Plot 1), sensitive (red) and resistant (blue) cell lines are heavily intertwined. After initial multi-modal feature fusion (Plot 2), discernible preliminary clusters form, though some overlap remains. Following complete end-to-end training (Plot 3), cell lines form two distinct clusters with sharp boundaries and enhanced internal compactness, achieving superior separation. This robust classification is primarily due to the Dual-Perturbation contrastive self-supervised learning framework, which enables the model to capture intrinsic discriminative patterns for drug response prediction and effectively separate response classes in the embedding space. These visualizations provide compelling evidence for the efficacy of the HGACL-DRP model design.

## 4. Conclusion and Outlook

The significant heterogeneity of cancer poses a substantial challenge to the realization of precision medicine, with the accurate prediction of patient drug response representing a critical bottleneck. The primary contribution of this research is the development of a high-performance computational tool tailored for drug response prediction. Its advantages are highlighted in several key aspects: First, the proposed attention-driven multi-modal data fusion and kernel extraction method provides an innovative solution for effectively integrating heterogeneous biological data. Second, the reliable neighbor filtering mechanism enables the construction of high-quality biological heterogeneous graphs with greater precision. Most importantly, the Dual-Perturbation contrastive learning framework, specifically designed for drug response prediction, significantly enhances the model’s generalization and discriminative capabilities in complex biological scenarios, thereby addressing the challenge of label scarcity in the biomedical domain through a novel strategy.

Although HGACL-DRP has achieved remarkable success, there remains considerable scope for further exploration and optimization. Future efforts will concentrate on integrating multidimensional biological information, such as proteomics data, and leveraging prior knowledge, including drug-target interactions and protein-protein interactions, to construct a more comprehensive biomedical knowledge graph. Simultaneously, enhancing the model’s interpretability to elucidate the underlying biological mechanisms of its predictions is crucial for gaining clinical trust. Ultimately, we aim to validate the model using patient-derived, real-world datasets to accelerate its translation into clinical applications.

## Data Availability

All data produced in the present work are contained in the manuscript

## Data and Code Availability

The data and code used are available at https://github.com/MingJin426/HGACL-DRP.

## Acknowledgments

Research on this work is partially supported by grants from the National Natural Science Foundation of China (No.62166028).

## Notes

### Competing Interest Statement

The authors have declared no competing interest.

